# The Impact of the November 2020 English National Lockdown on COVID-19 case counts

**DOI:** 10.1101/2021.01.03.21249169

**Authors:** Paul R Hunter, Julii Brainard, Alastair Grant

## Abstract

In the UK the epidemic of COVID-19 continues to pose a significant threat to public health. On the 14^th^ October the English government introduced a tier system for control of the epidemic but just 3 weeks later a National lockdown across all areas of England was implemented. When English areas emerged from Lockdown many were placed in different tiers (most typically moved up at least one tier). However, the effectiveness of the tier system has been challenged by the emergence of a new variant of SARS-CoV-2 which appears to be much more infectious. In addition, from early November a trial mass testing service was being run in Liverpool. We used publicly available data of daily cases by local authority (local government areas) and estimated the reproductive rate (R value) of the epidemic based on 7-day case numbers compared with the previous 7-day period. There was a clear surge in infections from a few days before to several days after the lockdown was implemented. But this surge was almost exclusively associated with Tier 1 and Tier 2 authorities. In Tier 3 authorities where hospitality venues were only allowed to operate as restaurants there was no such surge. After this initial surge, cases declined in all three tiers with the R value dropping to a mean of about 0.7 independent of tier. London, The South East and East of England Regions saw rising infection rates in the last week or so of lockdown primarily in children of secondary school age. We could find no obvious benefit of the trial mass screening programme in Liverpool city. We conclude that in Tiers 1 and 2 much of the beneficial impact of the national lockdown was lost probably because of the leak of its likely implementation five days early leading to increased socialising in these areas before the start of lockdown. We further conclude that given that the new variant is estimated to have an R value of between 0.39 and 0.93 greater than previous variants, any lockdown as strict as the November one would be insufficient to reverse the increase in infections by itself. The value of city-wide mass testing to control the epidemic remains uncertain.

## Introduction

At the time of writing SARS-CoV-2 has been reported as causing over 80 million cases and over 1.7 million deaths worldwide (https://www.worldometers.info/coronavirus/). In the UK total case numbers are approaching 2.4 million and deaths with COVID on the death certificate are almost 80,000 (https://coronavirus.data.gov.uk/). Until the widespread take up of a safe and effective vaccine control of the pandemic has relied on nonpharmaceutical interventions that may or may not be successful (Brauner *et al*. 2020, Hunter *et al*. 2020, Li et al. 2020). The United Kingdom has not had a unified approach to control of the epidemic with each of the four nations (England, Wales, Scotland and Northern Ireland each following their own strategies. England started to follow a more sub-regional approach, culminating on the 14^th^ October with a three tier system (Prime Minister’s Office 2020). The main differences between the tiers were around socialising and opening of hospitality venues such as pubs, bars and restaurants. In Tier 1 people were allowed to meet up in homes in groups or no more than six. Pubs and restaurants could remain open but only serve food or drink to people whilst seated. In Tier 2, people who were not in the same household or support bubble were prohibited for socialising in any indoor setting though could socialise outdoors in groups of no more than six people. As in Tier 1, pubs and restaurants in Tier 2 could remain open but only serve food or drink to people whilst seated. But only people from the same household or support bubble could socialise together in these pubs and restaurants. In Tier 3 people could not socialise indoors, in their own gardens or at most outdoor settings. Hospitality venues were only allowed to operate as restaurants.

However, in the last two days of October it was widely leaked that the English government was going to impose a new national lockdown (Blackall 2020). This lockdown eventually came into force on the 5th of November. During this new lockdown, people were instructed to stay at home except for specific purposes; non-essential retail was closed, but schools and universities remained open (Department of Health and Social Care 2020). The lockdown ended on the 2^nd^ December.

That the November lockdown would be imposed was leaked to news media just over 2 weeks after the start of the new tier system was implemented. The November lockdown started before an adequate assessment of the effectiveness of the tier system could be made. We undertook an analysis of the effectiveness of the tier system using case data available through 12 Nov 2020 (Hunter et al. 2020). To generalise, we found that in Tier 3 authorities there was a continuing decline in cases and that the mean R value was about 0.9 after 14 days in that tier. In Tier 2 the mean R value was about 1.0 and in Tier 1, the mean R was about 1.5. In Tier 1 local authorities were still experiencing exponential growth. In Tier 2 the epidemic was on average level and in Tier 3 the epidemic was declining. We concluded that the tier system could have controlled the epidemic but local authorities had not been placed into the most appropriate tiers or been moved into a higher tier quickly enough. We suggested that the faster decisions to move local authorities into higher tiers could be justified based on emerging evidence

Another observation was that around the start of the National lockdown on the 5^th^ November, case numbers started to increased. This seemed to relate to increased socialising following premature publicity about the November lockdown plans (Hunter et al. 2020). In this paper we continue our analyses from our initial tier paper to determine what value the national lockdown had on controlling the epidemic in England and particularly to investigate the impact of this pre-lockdown surge in infections. In addition we took the opportunity to determine whether there was any observable impact of the Liverpool mass testing trial on the trajectory of the epidemic in that city.

## Methods

We followed the same methodology as in our previous paper (Hunter et al. 2020). All data on daily numbers of new cases of COVID-19 were downloaded from the English Department of Health and social care daily COVID dashboard at https://coronavirus.data.gov.uk/details/download.

As we stated in our previous paper “For each local authority, an estimate of the effective reproductive number (R) for preceding days was obtained by summing all reports of new COVID infections for each day and the previous six days. This was then compared with the sum of new cases over the previous seven-day period. R was estimated using the equation given by Grant (2020) as

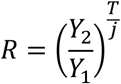

where Y_1_ and Y_2_ are the numbers of cases in two consecutive weeks, T is the generation time for the infection (here taken as 5 days Lauer et al. 2020) and j is the interval between the midpoints of the periods over which cases are counted (here 7 days). This estimator performs particularly well when R is close to 1 (Grant 2020).

We analysed data from all local authorities and also by subgroup based on whatever tier that the areawas allocated to on the last day of the first-tier system (i.e., 4^th^ November 2020).

Finally, we also took the opportunity to estimate the different epidemiology of the new variant strain that arose in London or the south east of England in the autumn of 2020 (Rambaut et al. 2020). Based on estimated proportion of the new cases that were due to the new strain in the weeks ending 18^th^ November and the 9^th^ December as presented at the Downing Street press conference of the 19^th^ December https://assets.publishing.service.gov.uk/government/uploads/system/uploads/attachment_data/file/946402/COVID-19_Press_Conference_Slides_-_Saturday_19_December.pdf and known total case reports from those regions for the same periods, we estimated the new number of cases of the new variant London, the South East and East of England Regions and compare those numbers with the number of non-new variant cases.

## Results

Data were obtained on 315 local authorities. The daily cases numbers for England as a whole are shown in Figure 1. It can be clearly seen that case numbers were increasing up to about 24^th^ October when there was a suggestion that the epidemic may have been plateauing. However, in the few days before the national lockdown case numbers started to rise again until the they reached a peak on about the 10^th^ November before declining until about the 30^th^ after which cases started to rise again.

**Figure 1.**
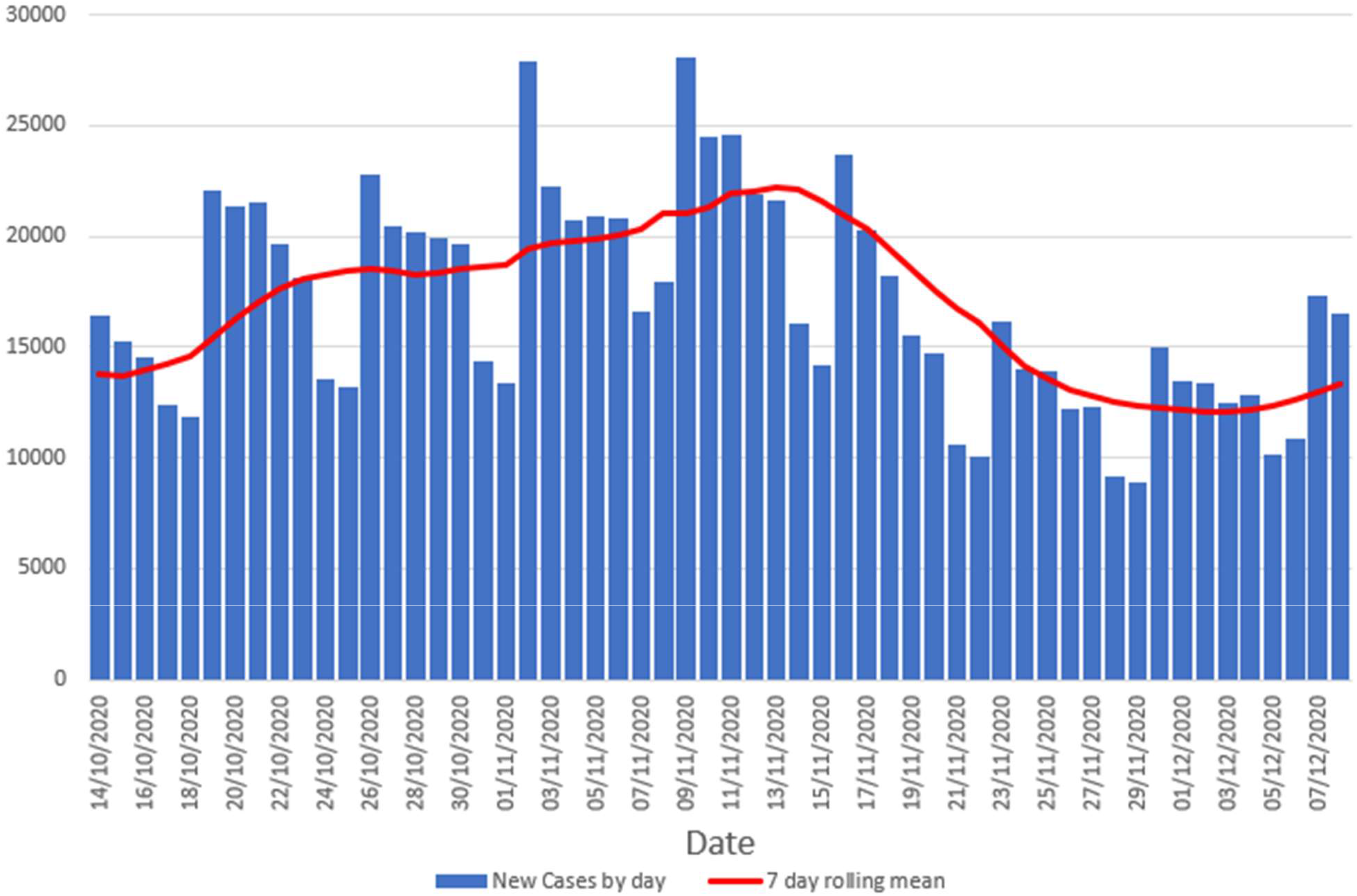
New case number in by specimen date for England.

But there was significant variation in the risk of COVID-19 cases across the UK. Figure 2 shows these same data but as rolling 7-day incidence per 100,000 people broken down by region and the associated R values. It is clear that for most regions, the R values were falling to about the 30^th^ October when the forthcoming National Lockdown was leaked to the press. After that the R values increased to peak on around the 14^th^ November before declining once more. However, about a week before the end of the National Lockdown (approximately 25^th^ November) the R value started to increase once more. For three regions, London, The South East and East of England R increased above 1. These changes in R were reflected in changes in the incidence with case numbers starting to increase even before the end of lockdown.

**Figure 2.**
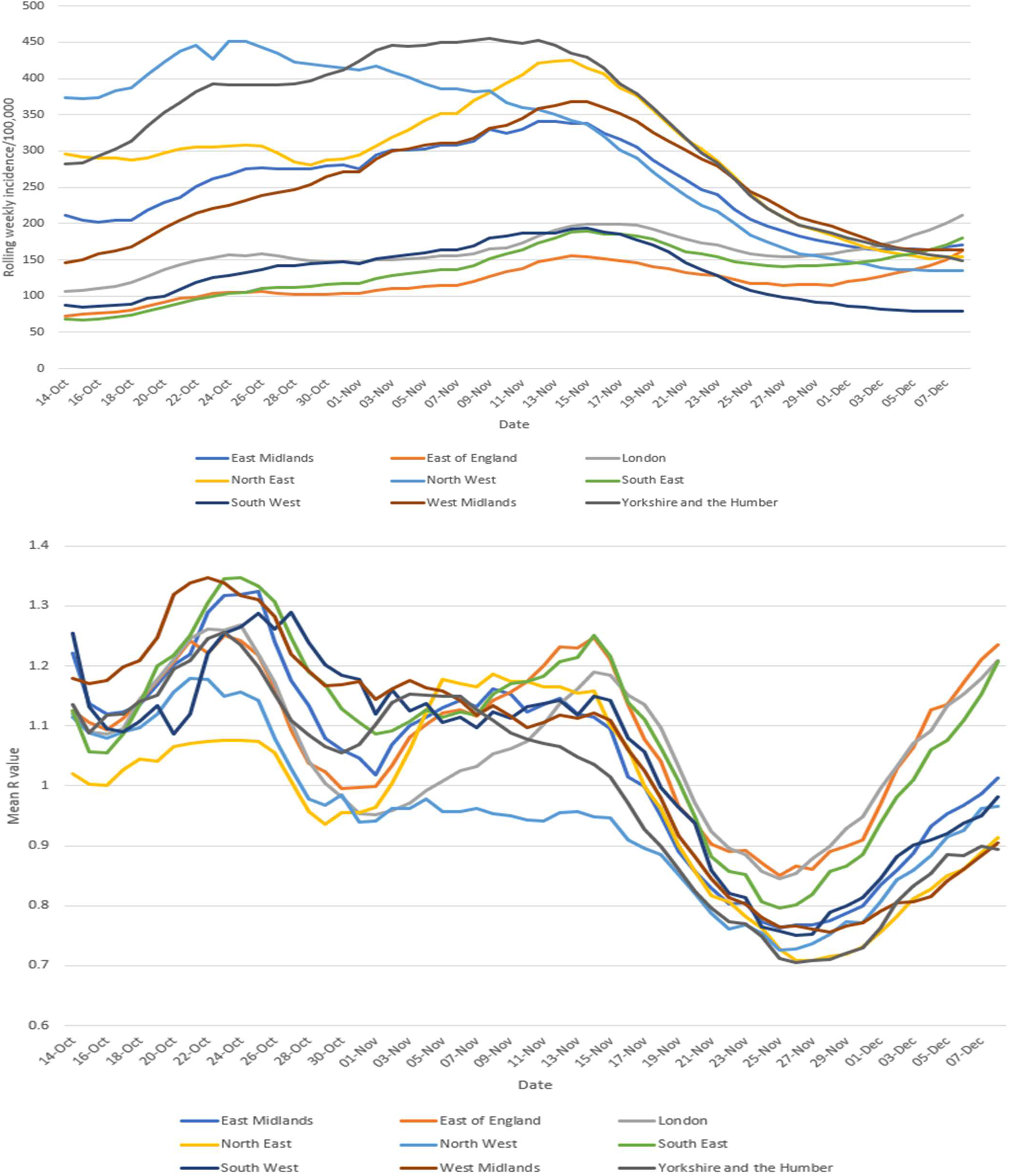
Rolling 7 day incidence by region and associated mean R values by region.

Figure 3 shows these same analyses but this time with incidence and R by which tier local authorities had been in on the 4^th^ November, the day before lockdown. There was a notable decline in R value during the first two weeks of the October tier system in authorities in all three tiers but after the 20^th^ October R increased in tiers one and two but not in Tier 3 to a peak around 13^th^ November. There was no such peak in R in Tier 3 authorities, instead we saw a steady decline up to about the 25^th^ November. Tiers 1 and 2 also declined up to the 25^th^ and on that date the R value was similar across the three tiers. Unfortunately, R stated to increase once more, most dramatically in authorities that had previously been in Tiers 1 and 2. Again changes in R were reflected in changing incidence. Local authorities that had previously been in Tier 3 had a much higher incidence at the start of lockdown but did not experience of surge of cases in early November. Tier 1 and 2 Authorities did see such a surge and by the end of Lockdown previously Tier 2 authorities were showing a higher rolling 7-day incidence than previously Tier 3 authorities

**Figure 3.**
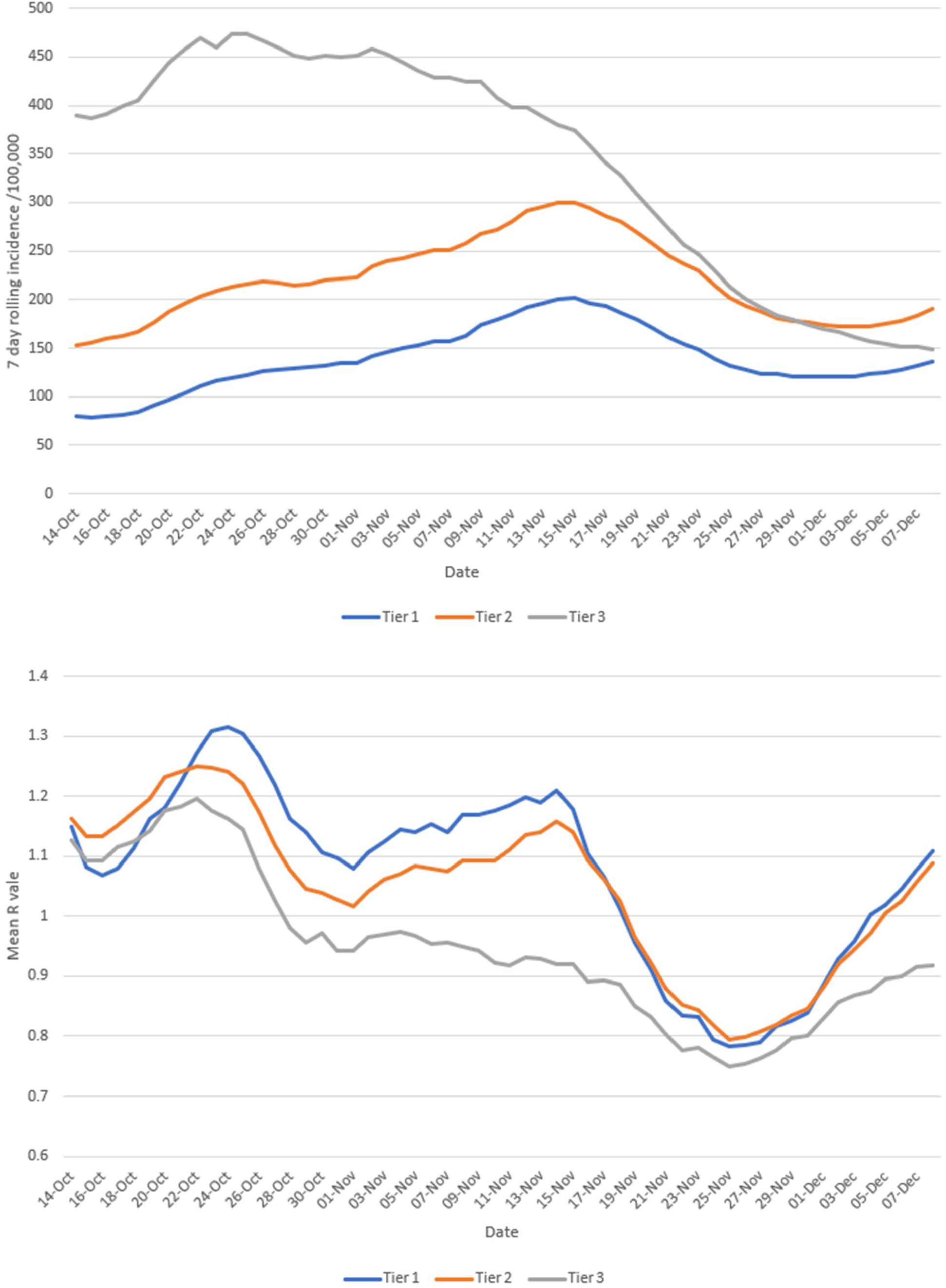
Rolling 7-day incidence by tier on day before lockdown and associated mean R values.

To investigate the early November and early December surges in infection further we present data on age specific rolling incidence as available on the DHSC COVID dashboard. Figures in the appendix show the rolling weekly incidence for each 5 year age group by region. In addition, we present the graph for the London Region in figure 4. It is notable that in general the ages groups between 30 to 59 show very similar patterns of incidence in the same region, showing a peak about the 14^th^ November before declining, with an increase in early December. The North West region, previously in Tier 3 did not show such a peak. The over 60 age groups were much less affected by the lockdown possibly reflecting higher adherence to social distancing and shielding in these groups prior to and continuing during lockdown. The most obvious temporal changes were in the 10 to 29 age groups. In almost all regions the early November surge in cases were most obvious in the 20 to 24 and 25 to 29 year age groups. But after lockdown the incidence dropped in these age groups. In several regions the 15 to 19-year age group had a similar temporal pattern to their older peers.

**Figure 4.**
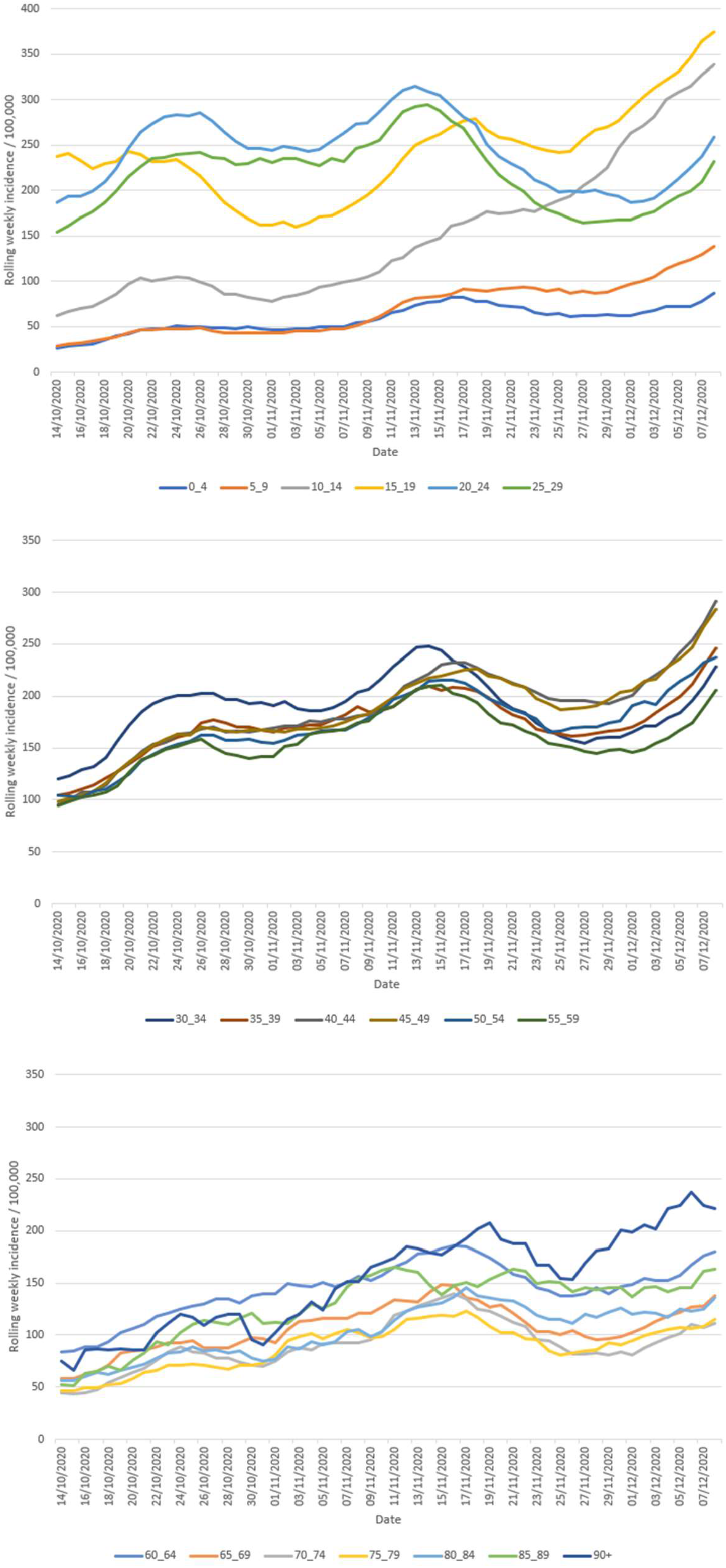
Age specific rolling weekly incidence per 100,000 – London Region.

The most striking observation was that in those regions that were seeing growth towards the end of the November lockdown (London, South East and East of England), there was an increase in cases in the 10 to 14 year age group, and to a lesser extent increases in the 15 to 19 age group. Especially in London it can be seen that case numbers in the 10 to 14 age groups grew steadily throughout the lockdown period and that case counts in the 15 to 19 age group also rose from about the 25^th^ November.

We also took the opportunity to see whether we could identify any impact of the mass testing being initially rolled out in Liverpool from 6^th^ November but extended throughout the month (Anon 2020). Figure 5 shows the daily estimates of R for Liverpool city, the mean of the R for other authorities in the Liverpool City area, excluding Liverpool city and the mean R for all other authorities in the North West region excluding Liverpool city area. It can be seen that there was a strong and temporary increase in R around the start of Lockdown in Liverpool, not obviously replicated in the wider region but similar to what was observed in Tier 1 and Tier 2 areas. Towards the end of the analysis period the R value in Liverpool city was very similar to that in other nearby authorities. Consequently, we were unable to identify any particular benefit of the Liverpool mass testing project on the overall trajectory of the epidemic in that city.

**Figure 5.**
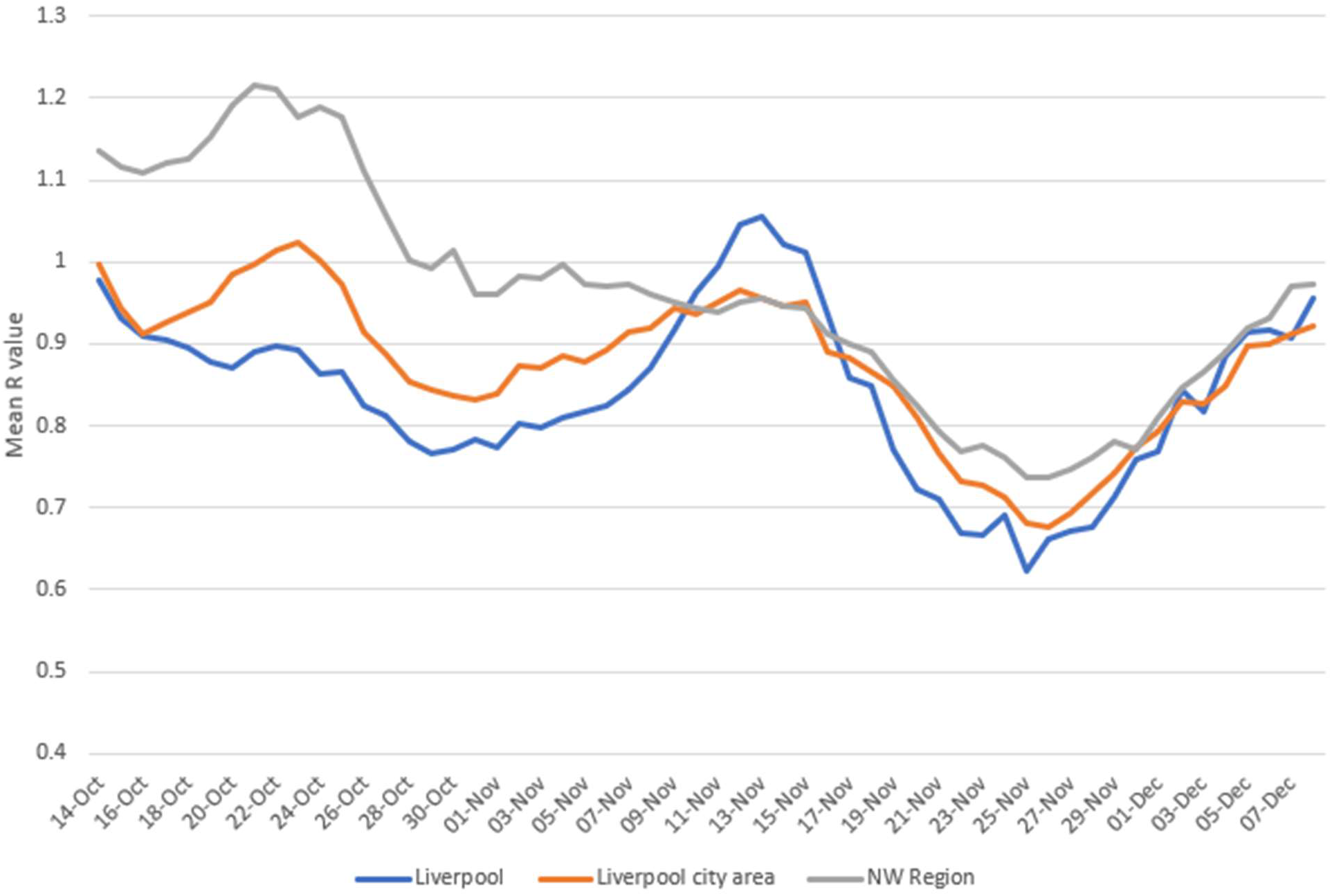
Daily estimate of R value for Liverpool city, the mean of the R for other authorities in the Liverpool City area, excluding Liverpool city and the mean R for all other authorities in the North West region excluding Liverpool city area.

**Figure 6.**
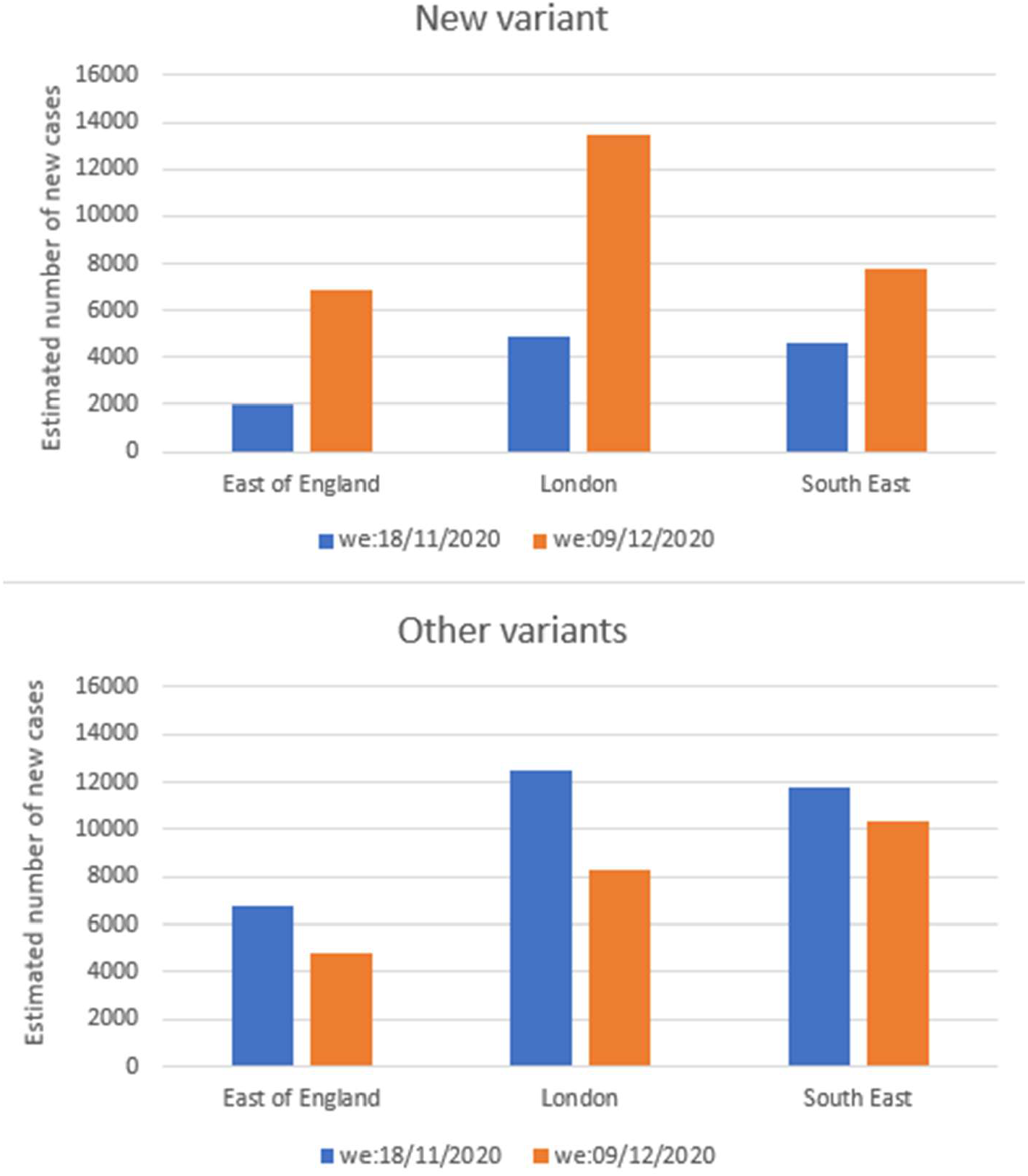
Estimated case numbers of a new variant of SARS-CoV-2 in three English Regions. Note: we=week ending.

In Figure 5 we show the estimated number of cases due to the new variant and all other variants. It can be seen that from mid-lockdown in November to the first week in December, most variants declined but the new variant increased quite dramatically during that same time. The estimated R value for the East of England, London and South East regions for the novel variant was 1.3, 1.3 and 1.1 respectively indicating growth in the epidemic of new variant cases. The equivalent R values for all other strains combined were 0.92, 0.91 and 0.97.

## Discussion

We undertook an analysis of new cases of COVID-19 by local authority in England from the 14^th^ of October to the 8^th^ of December. We found that for those authorities previously in Tier 3, the epidemic was already in decline, prior to the national lockdown but that the lockdown accelerated this decline. For authorities in Tiers 1 and 2 however, there was a marked increase in cases starting soon after the lockdown it was leaked that a lockdown was likely and continuing for up to two weeks after the start of the lockdown. It has been suggested that this surge of cases around the start of lockdown was due to increased socialising. Google mobility did detect an increase in visits to non-grocery retail and leisure venues just prior to the start of lockdown in England but not in Scotland (Google 2020). Given that this early lockdown spike did not occur in Tier 3, where the main difference was that pubs could only open if operating as a restaurant and the predominant age and that the spike was most obvious in the 20 to 24 and 25 to 29 age groups, a link to increased socialising in the days before lockdown is plausible. In any event, it would appear that the value of the lockdown in Tier 1 and 2 authorities was largely dissipated by this early November surge. We could find no clear evidence of any important impact of the mass testing intervention in Liverpool as a result of the mass testing pilot that was started in that city early in November.

The increase in cases that started during the last week of lockdown in London, The South East and East of England Regions is extremely worrying. The analyses presented here suggest that the infection started spreading initially in secondary school aged children especially in those aged 10 to 14 years old and soon drove increases in other age groups. However, more recent evidence of a novel and more infectious strain (B.1.1.7 lineage) in those same regions is another important explanation (Rambaut et al. 2020).

Whether the new tier system introduced on the 2^nd^ December at the end of the national lockdown in England would have been effective in the controlling the epidemic in England, we will probably never now know. It is likely that the new variant will become the dominant type over coming weeks. At best during the national lockdown we managed to reduce the mean of local R values to be about 0.7. All three tiers are likely to have been associated with mean R values somewhat greater than that. If the new variant does indeed add 0.4 or greater to the R value then it is anticipated that all areas will need to move ‘up’ at least one tier, in order to control the outbreak, at least until effective vaccination is widespread.

In conclusion, we have shown that the beneficial effects of the national lockdown in authorities in Tiers 1 and 2 were probably undermined because of increased socialising in the days before implementation. Leaked plans about the Lockdown before its implementation indirectly led to a surge in infections in the five day ‘notice’ period between public awareness and the start of Lockdown. In London, the South East and East of England regions cases started rising in the week before lockdown ended. The initial rise in these regions was first observed in children of secondary school age and especially the 10 to 14 year age group, though towards the end of lockdown increases were seen in older age groups in these regions. We now know that that much of this increase was associated with a new variant that has higher infectivity (NERVTAG 2020). From our analysis it is doubtful that even a return to the November lockdown conditions would be sufficient to control spread of the new variant.

## Data Availability

All data used in this paper is publicly available

https://coronavirus.data.gov.uk/details/download

## Appendix

**Figure S1.**
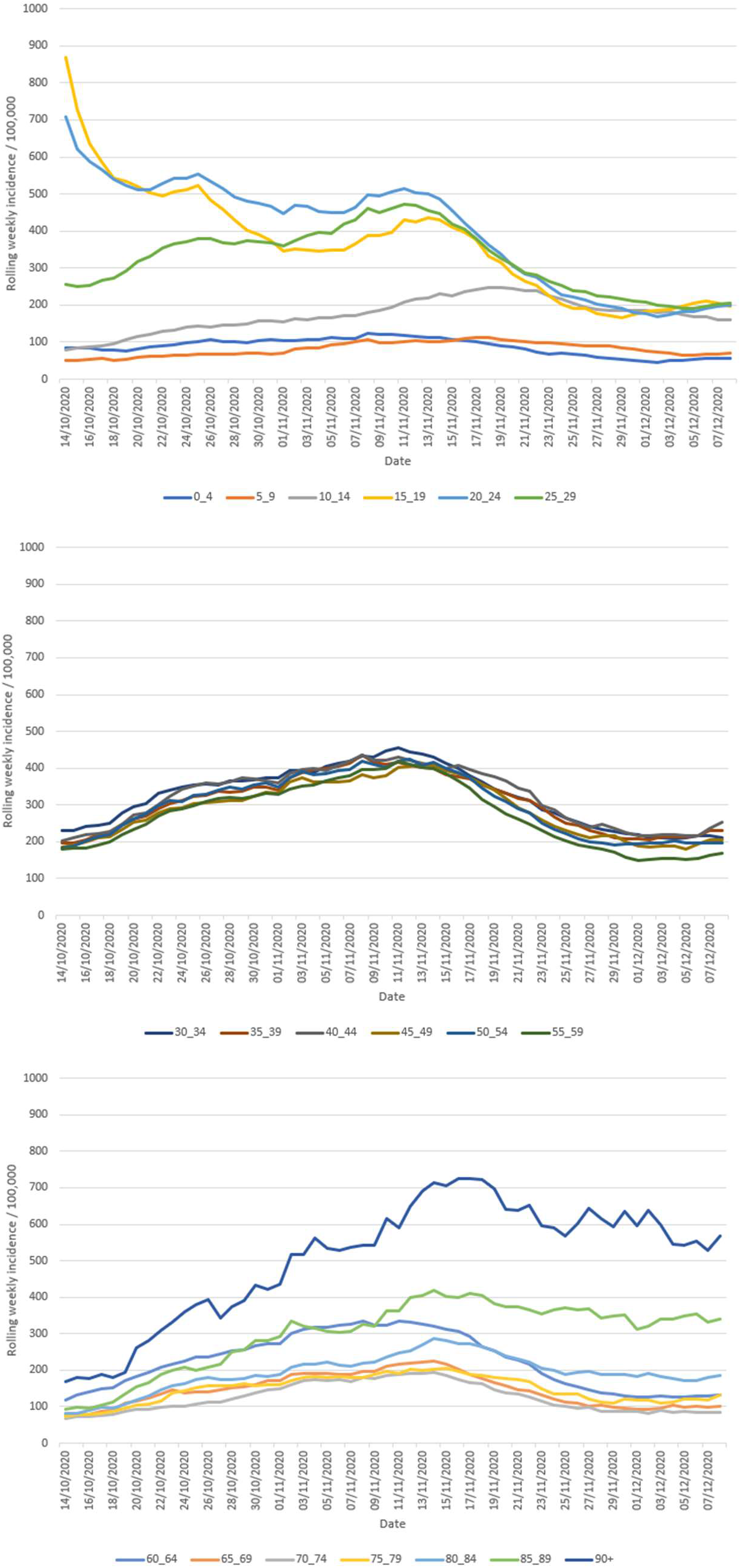
Ages specific rolling weekly incidence per 100,000 - East Midlands Region.

**Figure S2.**
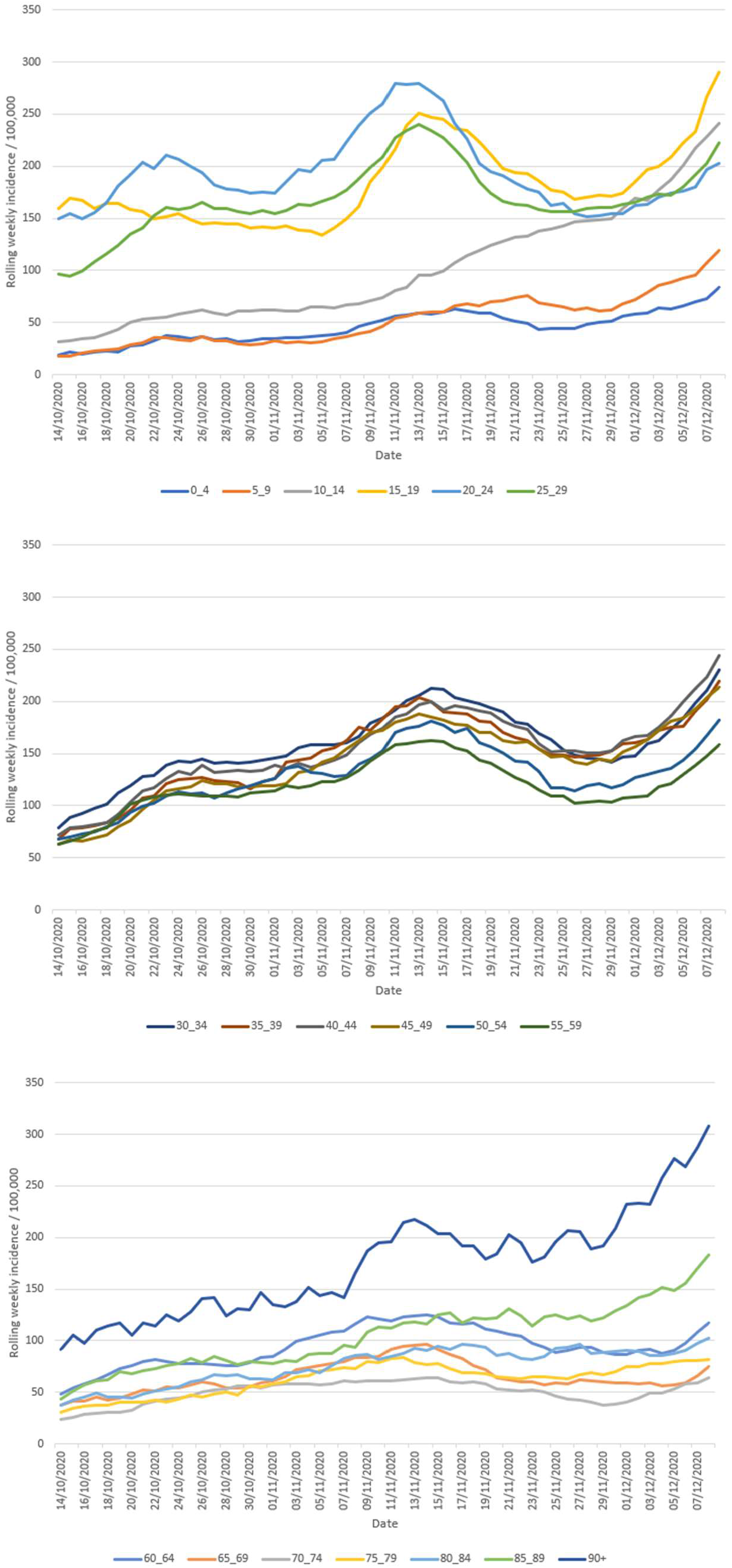
Ages specific rolling weekly incidence per 100,000 - East of England Region.

**Figure S3.**
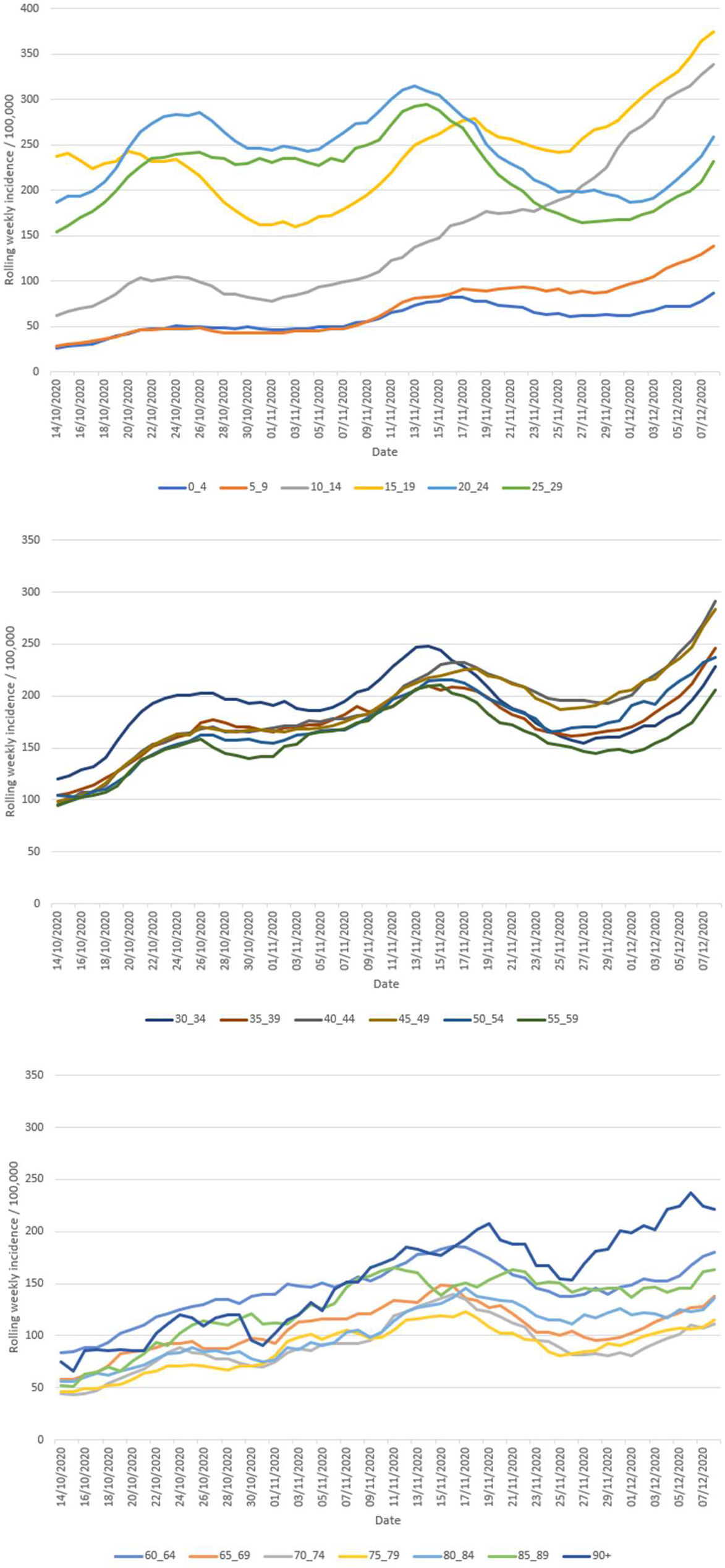
Ages specific rolling weekly incidence per 100,000 – London Region.

**Figure S4.**
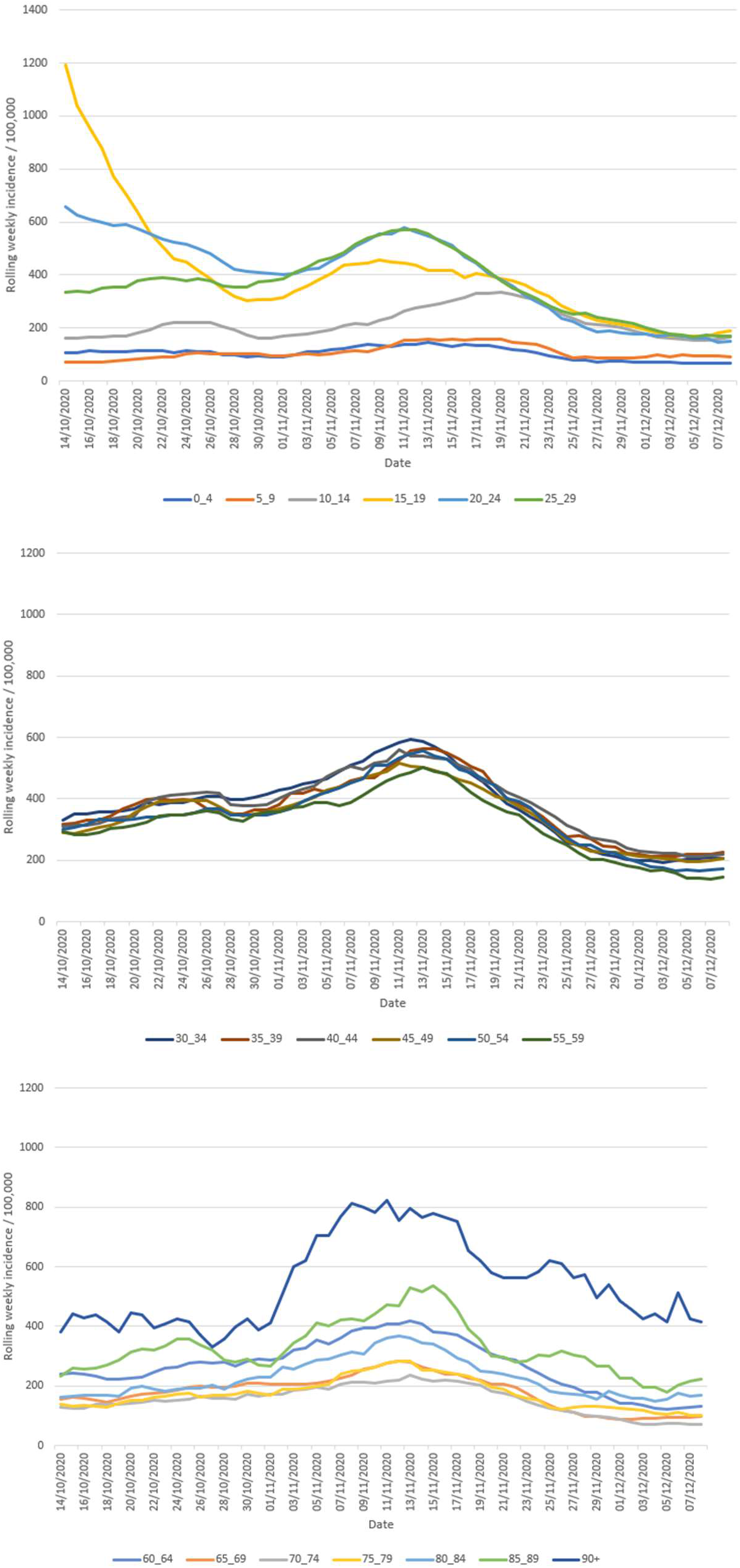
Ages specific rolling weekly incidence per 100,000 – North East Region.

**Figure S5.**
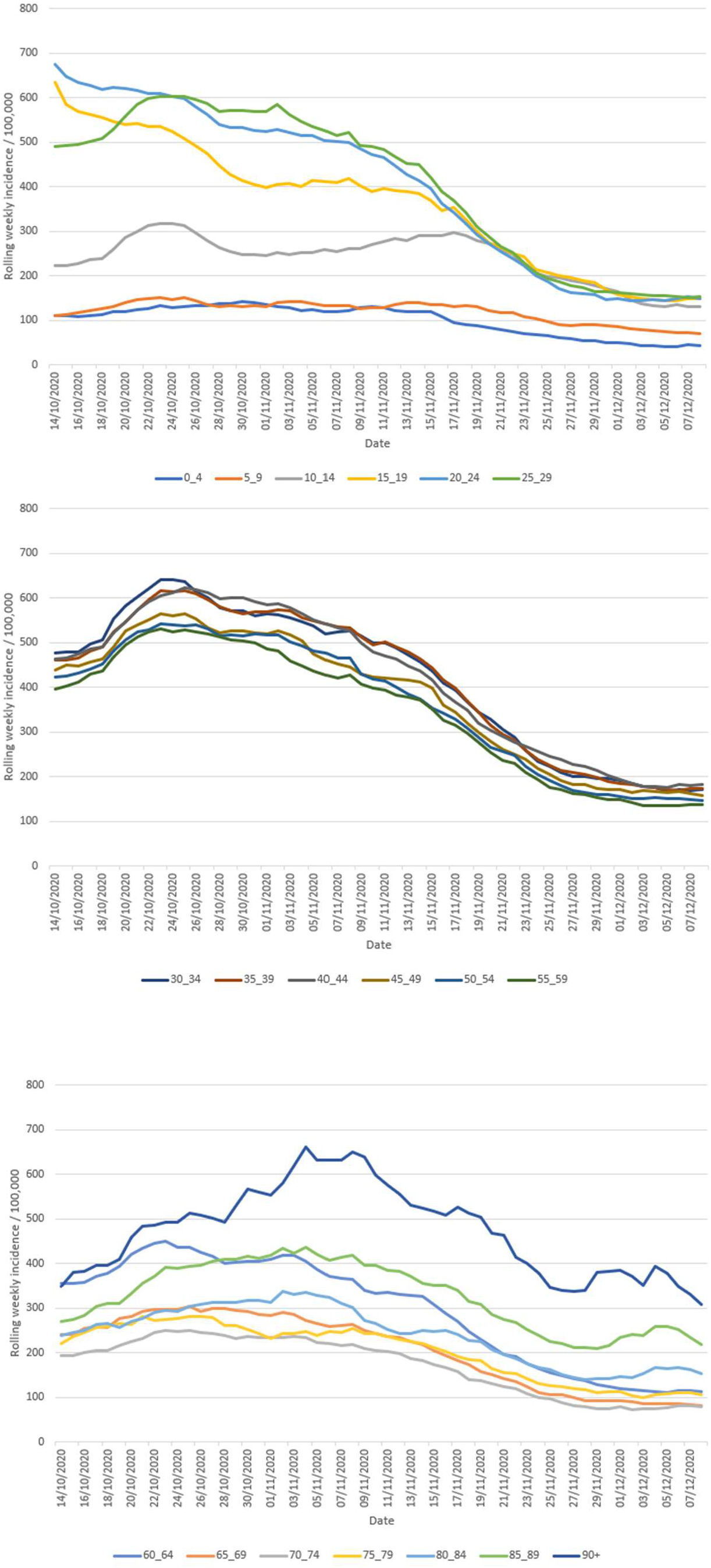
Ages specific rolling weekly incidence per 100,000 – North West Region.

**Figure S6.**
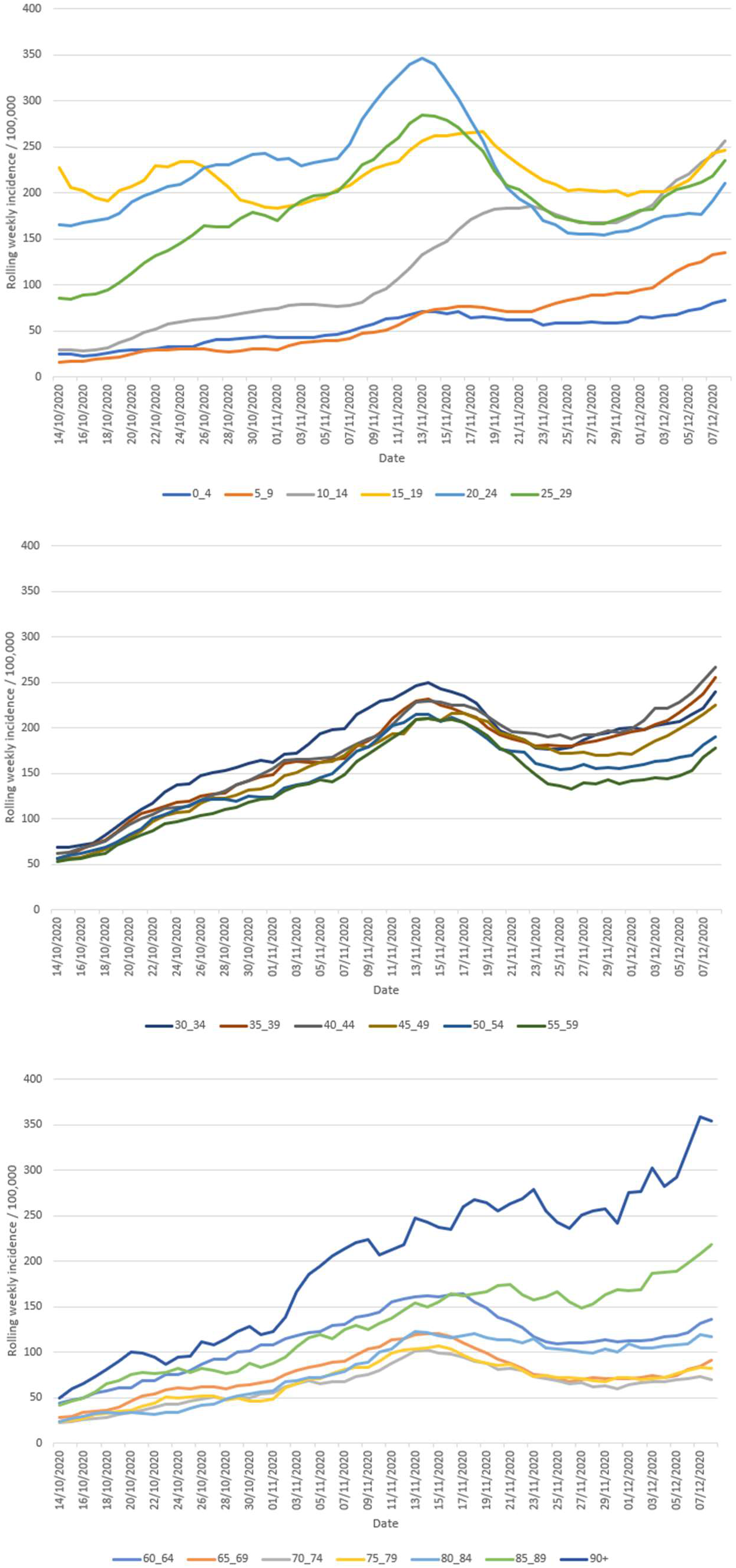
Ages specific rolling weekly incidence per 100,000 – South East Region.

**Figure S7.**
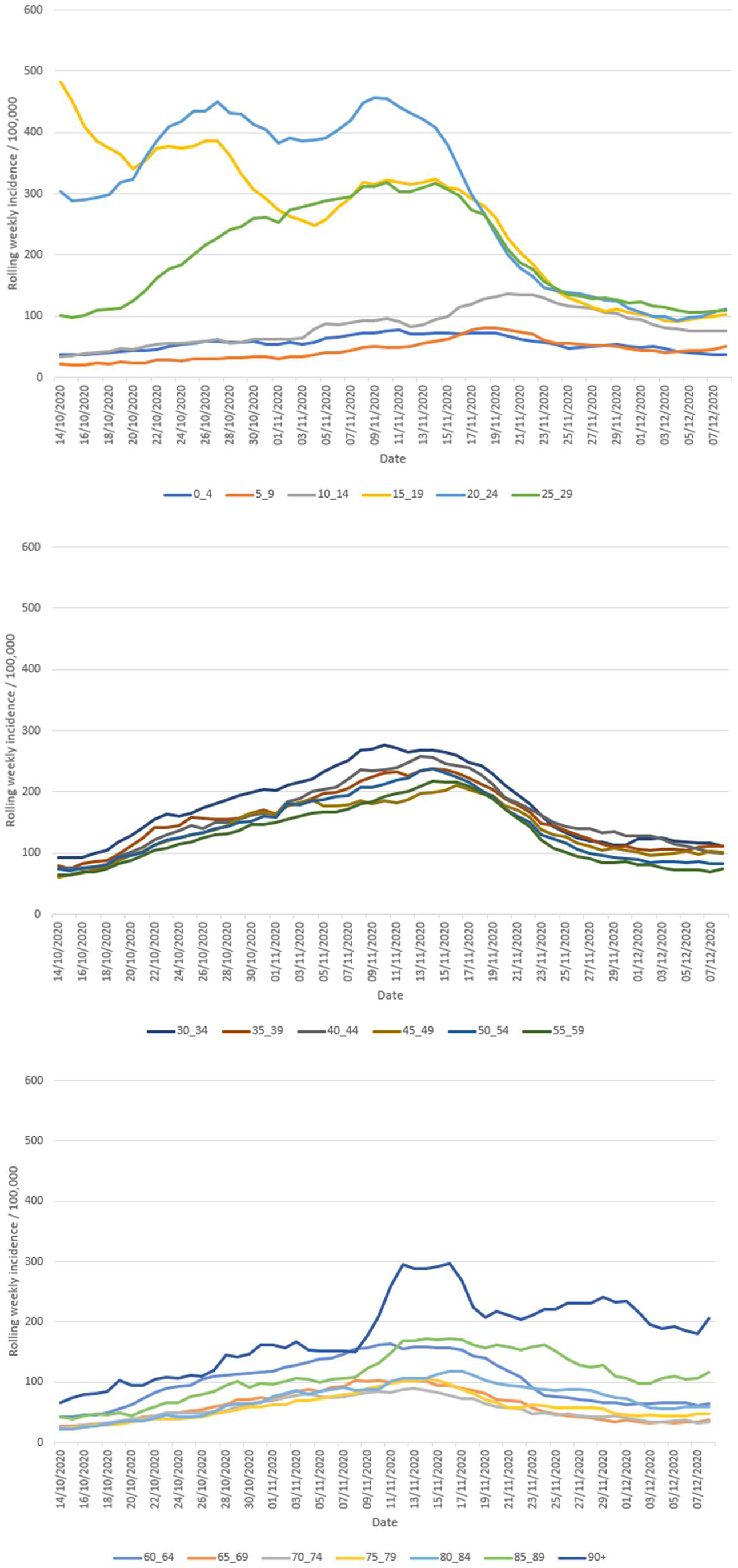
Ages specific rolling weekly incidence per 100,000 – South West Region.

**Figure S8.**
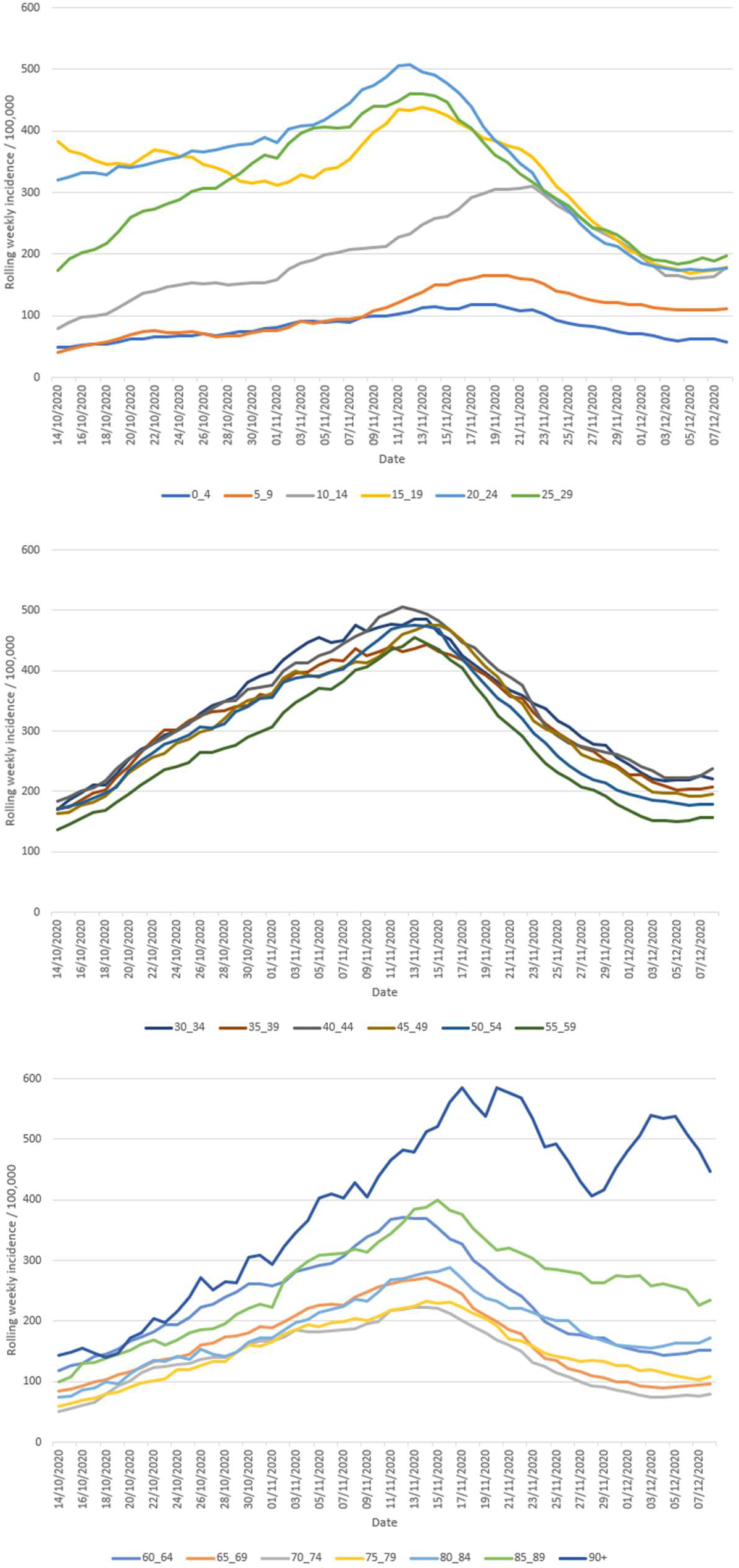
Ages specific rolling weekly incidence per 100,000 –West Midlands Region.

**Figure S9.**
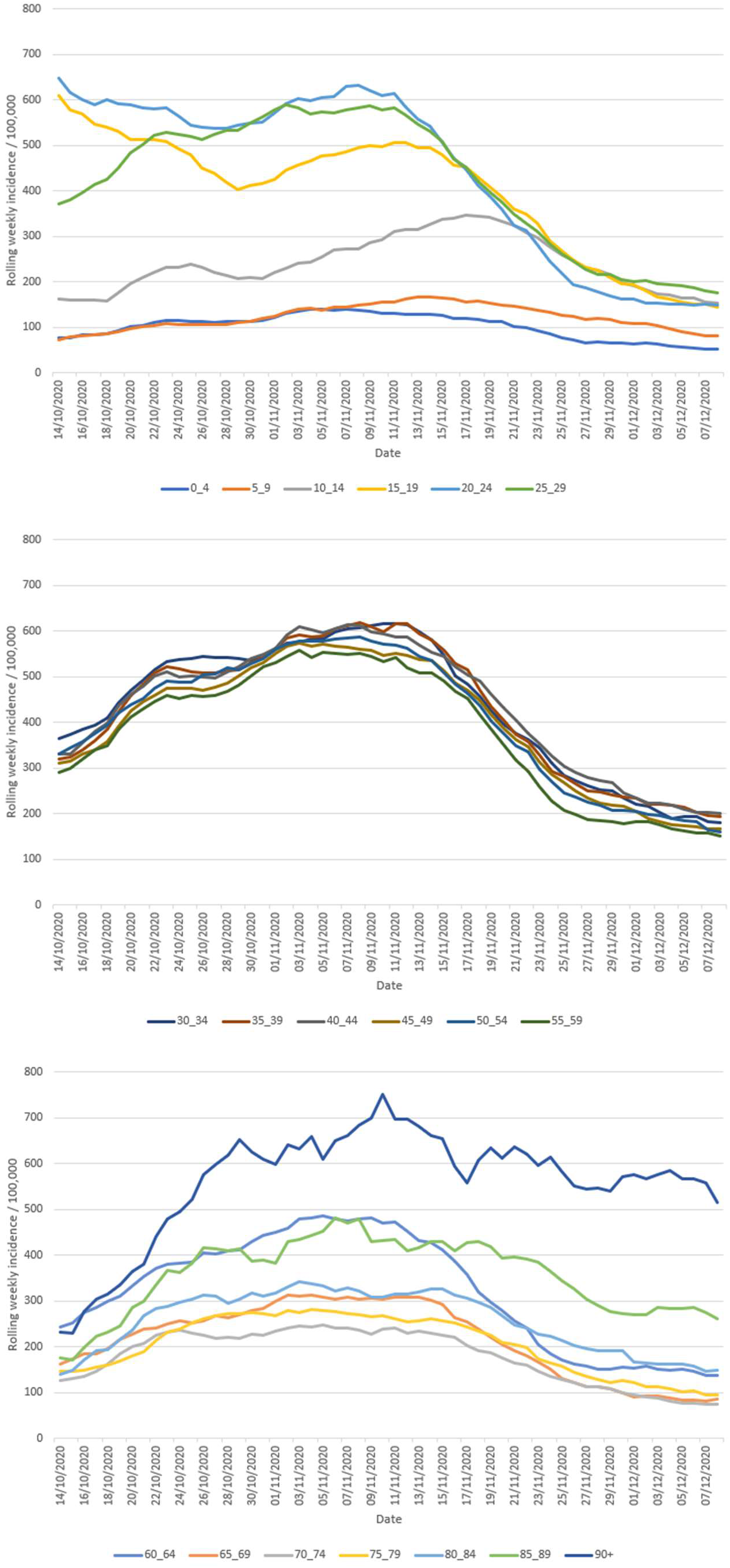
Ages specific rolling weekly incidence per 100,000 –Yorkshire and The Humber Region.

